# Safety and immunogenicity of VLPCOV-02, a SARS-CoV-2 self-amplifying RNA vaccine with a modified base, 5-methylcytosine, in healthy individuals

**DOI:** 10.1101/2023.09.04.23294493

**Authors:** Masayuki Aboshi, Kenta Matsuda, Daisuke Kawakami, Kaoru Kono, Yoko Kazami, Takashi Sekida, Mai Komori, Amber L. Morey, Shigeru Suga, Jonathan F. Smith, Takasuke Fukuhara, Yasumasa Iwatani, Takuya Yamamoto, Nobuaki Sato, Wataru Akahata

**Affiliations:** VLP Therapeutics Japan, Inc., 1-16-4 Nishi-Shinbashi, Minato-ku, Tokyo 105-0003, Japan; VLP Therapeutics, Inc., Gaithersburg, MD 20878, USA; National Hospital Organization, Mie National Hospital, Tsu, Mie 514-0125, Japan; Department of Microbiology and Immunology, Faculty of Medicine, Hokkaido University, Sapporo, Hokkaido 060-0815, Japan; Laboratory of Virus Control, Research Institute for Microbial Diseases, Osaka University, Suita, Osaka 565-0871, Japan; Clinical Research Center, National Hospital Organization Nagoya Medical Center, Nagoya, Aichi 460-0001, Japan; Division of Basic Medicine, Nagoya University Graduate School of Medicine, Nagoya, Aichi 466-8550, Japan; Laboratory of Precision Immunology, Center for Intractable Diseases and ImmunoGenomics, National Institutes of Biomedical Innovation, Health and Nutrition, Ibaraki, Osaka 567-0085, Japan

**Keywords:** SARS-CoV-2, COVID-19, self-amplifying RNA, nucleoside-modified base, 5-methylcytosine, pan HLA DR-binding epitope sequence, booster vaccine, safety, immunogenicity

## Abstract

Continuing emergence of variants of concern resulting in reduced SARS-CoV-2 vaccine efficacy necessitates additional prevention strategies. The structure of VLPCOV-01, a lipid nanoparticle-encapsulated, self-amplifying RNA COVID-19 vaccine with a comparable immune response to BNT162b2, was revised by incorporating a modified base, 5-methylcytosine, to reduce reactogenicity, and an updated receptor-binding domain derived from Brazil (gamma) variant. Interim analyses of a phase 1 dose-escalation booster vaccination study with the resulting construct, VLPCOV-02, in healthy, previously vaccinated Japanese individuals (N=96) are reported (jRCT2051230005). A dose-related increase in solicited local and systemic adverse events was observed, which were generally rated mild or moderate. The most commonly occurring events were tenderness, pain, fatigue, and myalgia. Serum SARS-CoV-2 immunoglobulin titers increased during the 4 weeks post-immunization. VLPCOV-02 demonstrated a favorable safety profile compared with VLPCOV-01, with a lower frequency of adverse events and fewer fever events at an equivalent dose. These findings support further study of VLPCOV-02.

## INTRODUCTION

The emergence and spread of severe acute respiratory syndrome coronavirus 2 (SARS-CoV-2) and the resultant coronavirus disease 2019 (COVID-19) pandemic has resulted in approximately 768 million cases and almost 7 million deaths globally over the past 4 years.^1^ The pandemic triggered a rapid development and rollout of SARS-CoV-2 vaccines that mitigated the impact of the COVID-19 pandemic, particularly in terms of severe infection and death.^2,3^ At the time of writing, the World Health Organization has approved 11 vaccines developed on four distinct platforms (inactivated virus, mRNA, adenovirus vector, and protein subunit) for use worldwide.^4^ However, providing protection against the virus is challenging due to the inevitable and continuous emergence of new variants, as well as the waning vaccine efficacy among previously vaccinated individuals.^5,6^ Vaccine efficacy can be improved by booster vaccination, which can be further enhanced if individuals receive a vaccine that is different to the original vaccine series received.^7^ The optimum regimen for booster vaccinations has yet to be established and, as with the primary vaccination, the additional protection wanes over time.^3,8^ This highlights the need for alternative vaccine strategies for the prevention of COVID-19.

A lipid nanoparticle (LNP)-encapsulated, self-amplifying RNA (saRNA) vaccine platform has been developed with a replaceable antigenic domain that allows expedited development of vaccines to address emerging variants of concern. These saRNA vaccines require lower doses than their non-amplifying mRNA counterparts.^9,10^ A novel COVID-19 vaccine, VLPCOV-01, was developed using the LNP-encapsulated saRNA platform derived from an alphavirus amplification system that encodes replicase and transcriptase functions comprising the alphavirus nonstructural proteins (nsp1-4) from the attenuated TC-83 strain of Venezuelan equine encephalitis virus (VEEV), and which expresses a membrane-anchored receptor-binding domain (RBD).^11,12^ When delivered as a booster vaccine in the phase 1 VLPCOV-01-0102 clinical trial, VLPCOV-01 induced strong and persistent immune responses in both non-elderly and elderly healthy participants who had received primary vaccination with BNT162b2; neutralizing antibody titers to Wuhan, Delta, and Omicron variants of SARS-CoV-2 were observed.^11^ The immune response achieved was comparable to that observed with BNT162b2 and with a similar safety profile, but at one-tenth of the vaccine dose.

VLPCOV-02 is a newly developed vaccine that is currently under investigation as a booster for individuals who have been previously vaccinated against and/or infected with SARS-CoV-2. This vaccine included several modifications of VLPCOV-01 that were designed to improve its safety profile and efficacy, and to confer broader protection. The immunogenicity per dose was increased by improving the integrity and purity of the RNA,^13^ and the pan HLA DR-binding epitope (PADRE) sequence, a short stretch of peptide that binds to most common human leukocyte antigen–DR isotypes,^14^ was added to broaden the vaccine response. Most importantly, a nucleoside-modified base, 5-methylcytosine (5-MeC) was incorporated to reduce innate responses, resulting in reduced reactogenicity.^15,16^ N1-Methylpseudouridine was utilized in existing mRNA vaccines to reduce the innate responses;^17,18^ however, N1-methylpseudouridine appears to inhibit the VEEV RNA-dependent RNA polymerase or transcriptase function. Therefore, innate responses were minimized in VLPCOV-02 by incorporation of 5-MeC, an approach supported by both preclinical *in vitro* and animal model observations (manuscript in preparation). The resulting modified product, VLPCOV-02, is currently being studied as a booster vaccine in a clinical trial setting in Japan.

A phase 1/2 study, VLPCOV-02-0102, is under way to evaluate the safety, tolerability, immunogenicity, and optimal booster dose of VLPCOV-02 in healthy Japanese adults who have received prior immunization with an approved SARS-CoV-2 vaccine. The interim analyses of the phase 1 dose-escalation study, including initial safety, tolerability, and immunogenicity findings, are reported here.

## RESULTS

### Participant disposition and baseline characteristics

Between April 10, 2023, and May 15, 2023, 174 individuals were screened, among whom 78 were excluded from involvement (due to failure to meet ≥1 eligibility criterion [n = 23], withdrawal of consent [n = 10], or the planned population for each dose group was reached [n = 45]). Thus, 96 participants were enrolled (48 in each of the non-elderly and elderly cohorts) and received a single booster vaccination with VLPCOV-02 at one of four doses, 1, 3, 7.5, or 15 μg (n = 12 participants per dose level in both cohorts) (**Figure 1** and **Figure S1**). At the data cutoff date (June 18, 2023), all 96 participants who received injections remained on study.

**Figure 1.**
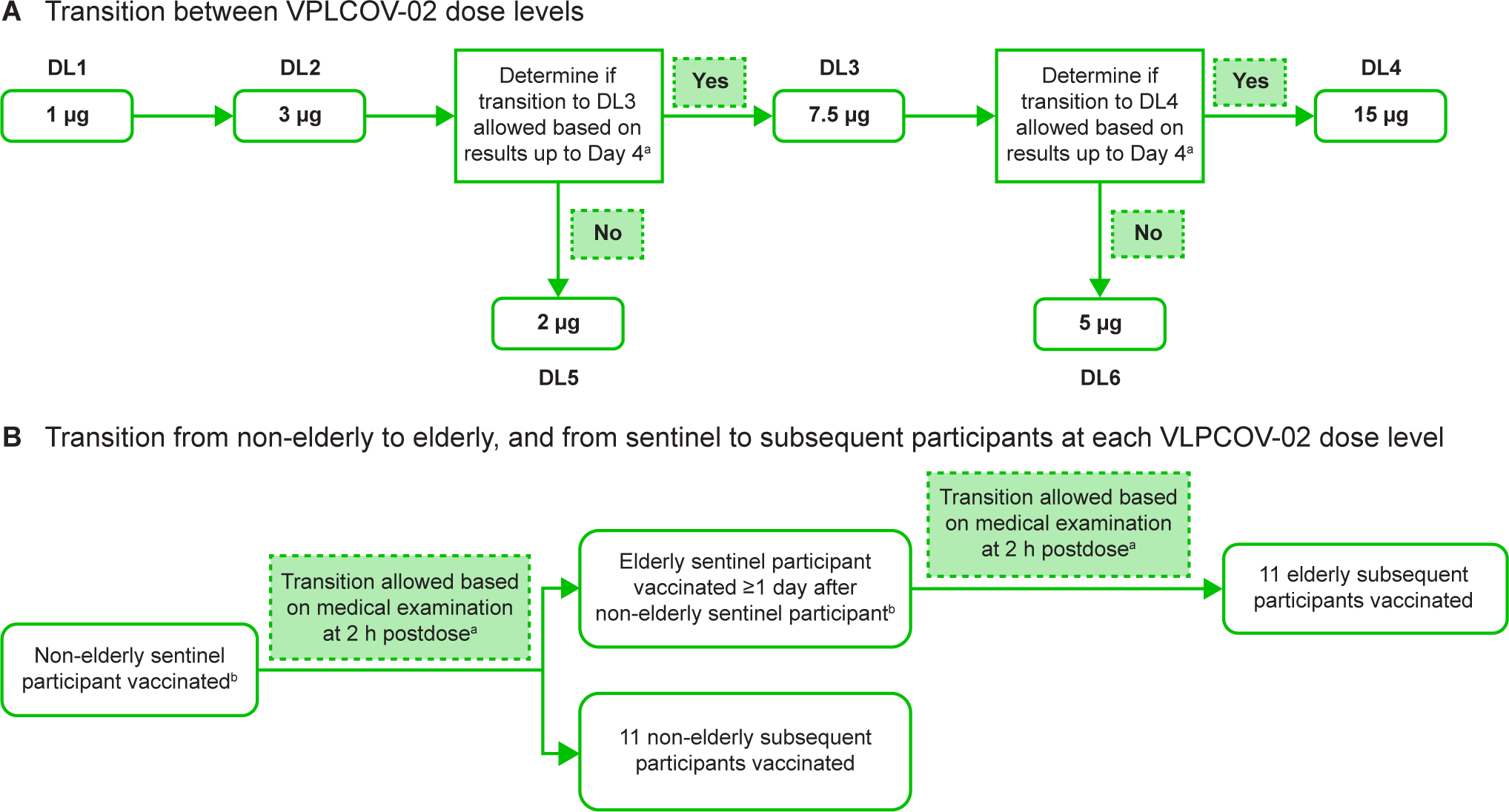
VLPCOV-02-0102 study design. **(A) Flow of transition between VLPCOV-02 dose levels. (B) Flow of transition from non-elderly to elderly participants, and from the sentinel to subsequent participants at each dose level.** ^a^The protocol prespecified discussion criteria for allowing transition to the next VLPCOV-02 dose level (DL) were: (1) the sentinel participant developed an acute symptom requiring emergent measures; (2) a participant in either cohort at the current DL developed a solicited grade 4 AE (according to the relevant United States Food and Drug Administration (FDA) Guidance for Industry^19^); (3) a participant in either cohort of the current DL developed a grade ≥3 AE (excluding solicited AEs) according to CTCAE v5.0; (4) ≥50% of participants in either cohort of the current DL developed solicited grade ≥3 AEs (according to the relevant FDA Guidance for Industry^19^) or grade ≥2 AEs according to CTCAE v5.0; (5) the principal investigator determined it necessary (even in the absence of criteria 1–4). ^b^In the case of transition to DL5 or DL6, no sentinel participant was required. AE, adverse event; CTCAE v5.0, Common Terminology Criteria for Adverse Events version 5.0; DL(n), dose level n.

The total study population included 51 (53.1%) females (n = 29 [56.9%] and n = 22 [43.1%] in the non-elderly and elderly cohorts, respectively) and was predominantly negative for history of COVID-19 infection (n = 73 [76.0%]: non-elderly, n = 35 [72.9%]; elderly, n = 38 [79.2%]) and for anti–N-protein antibodies (n = 84 [87.5%]: non-elderly, n = 43 [89.6%]; elderly, n = 41 [85.4]). The baseline characteristics of the study participants were generally comparable across the dose levels for both the non-elderly and elderly cohorts (**Table 1**). The non-elderly cohort (n = 48) had an overall age of 52.5 ± 7.1 years (mean ± SD; range 35–64 years), and that of the elderly cohort (n = 48) was 68.2 ± 2.7 years (range, 65–75 years). The non-elderly cohort was less intensively vaccinated than the elderly cohort, with 13 (27.1%) vs. 34 (70.8%) having received ≥4 prior SARS-CoV-2 vaccinations. One (2.1%) participant in the non-elderly cohort who received 3 µg VLPCOV-02 had received five prior SARS-CoV-2 vaccinations, compared with nine (18.8%) in the elderly cohort, one of whom received 3 µg VLPCOV-02 and eight received 15 µg VLPCOV-02. Mean time since the last vaccination was longer for the non-elderly cohort (range across dose levels: 11.8–13.0 months vs. 6.2–8.7 months for the elderly cohort).

**Table 1.** Baseline clinical and demographic characteristics of Japanese participants who received booster vaccination with VLPCOV-02 by dose and age cohort.

| Characteristic | VLPCOV-02 1 µg |  | VLPCOV-02 3 µg |  | VLPCOV-02 7.5 µg |  | VLPCOV-02 15 µg |  | Total population |  |
| --- | --- | --- | --- | --- | --- | --- | --- | --- | --- | --- |
|  | Non-elderly<br>n = 12 | Elderly<br>n = 12 | Non-elderly<br>n = 12 | Elderly<br>n = 12 | Non-elderly<br>n = 12 | Elderly<br>n = 12 | Non-elderly<br>n = 12 | Elderly<br>n = 12 | Non-elderly<br>n = 48 | Elderly<br>n = 48 |
| **Age (years)** |  |  |  |  |  |  |  |  |  |  |
| Mean ± SD | 52.3 ± 6.39 | 67.5 ± 2.88 | 52.8 ± 6.93 | 68.1 ± 2.31 | 50.8 ± 7.31 | 68.7 ± 3.28 | 54.0 ± 8.32 | 68.6 ± 2.68 | 52.5±7.1 | 68.2±2.8 |
| Median<br>(range) | 52.0<br>(40–64) | 67.0<br>(65–73) | 54.0<br>(35–61) | 68.0<br>(65–71) | 52.0<br>(36–63) | 68.0<br>(65–75) | 57.0<br>(36–64) | 68.0<br>(66–74) | 53.0<br>(35–64) | 67.5<br>(65–75) |
| **Sex, n (%)** |  |  |  |  |  |  |  |  |  |  |
| Male | 6 (50.0) | 5 (41.7) | 4 (33.3) | 7 (58.3) | 5 (41.7) | 6 (50.0) | 4 (33.3) | 8 (66.7) | 19 (39.6) | 26 (54.2) |
| Female | 6 (50.0) | 7 (58.3) | 8 (66.7) | 5 (41.7) | 7 (58.3) | 6 (50.0) | 8 (66.7) | 4 (33.3) | 29 (60.4) | 22 (45.8) |
| **History of COVID-19 infection, n (%)** |  |  |  |  |  |  |  |  |  |  |
| Yes | 2 (16.7) | 1 (8.3) | 4 (33.3) | 4 (33.3) | 5 (41.7) | 2 (16.7) | 2 (16.7) | 3 (25.0) | 13 (27.1) | 10 (20.8) |
| No | 10 (83.3) | 11 (91.7) | 8 (66.7) | 8 (66.7) | 7 (58.3) | 10 (83.3) | 10 (83.3) | 9 (75.0) | 35 (72.9) | 38 (79.2) |
| **Anti-N-protein antibody, n (%)** |  |  |  |  |  |  |  |  |  |  |
| Positive | 1 (8.3) | 2 (16.7) | 3 (25.0) | 2 (16.7) | 0 (0) | 1 (8.3) | 1 (8.3) | 2 (16.7) | 5 (10.4) | 7 (14.6) |
| Negative | 11 (91.7) | 10 (83.3) | 9 (75.0) | 10 (83.3) | 12 (100) | 11 (91.7) | 11 (91.7) | 10 (83.3) | 43 (89.6) | 41 (85.4) |
| **Baseline anti-RBD antibody titer<sup>a</sup>** |  |  |  |  |  |  |  |  |  |  |
| Geometric mean | 5260.6 | 10,099.2 | 13,066.9 | 13,188.8 | 13,130.6 | 12,771.3 | 12,660.6 | 21,877.2 | 10,339.2 | 13,889.3 |
| Min, Max | 1730,<br>51,000 | 732,<br>≥80,000 | 835,<br>62,900 | 1330,<br>≥80,000 | 1260,<br>42,800 | 1860,<br>58,800 | 4060,<br>66,800 | 2780,<br>≥80,000 | 835,<br>66,800 | 732,<br>≥80,000 |
| **Number of prior SARS-CoV-2 vaccines, n (%)<sup>b</sup>** |  |  |  |  |  |  |  |  |  |  |
| 2 | 1 (8.3) | 1 (8.3) | 0 (0) | 0 (0) | 3 (25.0) | 1 (8.3) | 2 (16.7) | 0 (0) | 6 (12.5) | 2 (4.2) |
| 3 | 9 (75.0) | 4 (33.3) | 9 (75.0) | 4 (33.3) | 6 (50.0) | 2 (16.7) | 5 (41.7) | 2 (16.7) | 29 (60.4) | 12 (25.0) |
| 4 | 2 (16.7) | 7 (58.3) | 3 (25.0) | 8 (66.7) | 3 (25.0) | 8 (66.7) | 4 (33.3) | 2 (16.7) | 12 (25.0) | 25 (52.1) |
| 5 | 0 (0) | 0 (0) | 0 (0) | 0 (0) | 0 (0) | 1 (8.3) | 1 (8.3) | 8 (66.7) | 1 (2.1) | 9 (18.8) |
| **Months since last vaccine** |  |  |  |  |  |  |  |  |  |  |
| Median | 12.1 | 8.1 | 12.3 | 8.7 | 11.8 | 8.2 | 13.0 | 6.2 | 12.2 | 7.8 |
| Range | 6.1–19.5 | 6.0–18.6 | 6.1–13.4 | 6.0–16.3 | 6.0–21.0 | 6.1–22.0 | 6.0–20.5 | 6.0–14.9 | 6.0–21.0 | 6.0–22.0 |

<sup>a</sup>The maximum titer measurable was 80,000; any values ≥80,000 were treated as 80,000 for the purposes of calculating the geometric mean value. <sup>b</sup>All participants had received at least a two-dose primary vaccination series with the same COVID-19 uridine-modified RNA vaccine or a booster inoculation with an mRNA vaccine (any kind, any number, and including bivalent mRNA vaccines) ≥6 months previously.
COVID-19, coronavirus disease 2019; SARS-CoV-2, severe acute respiratory syndrome coronavirus 2; SD, standard deviation.

### Safety

Following booster vaccination with VLPCOV-02, no serious adverse events (AEs) were reported, regardless of age or VLPCOV-02 dose received, and no participants had discontinued by the 4-week analysis time point. The majority of all solicited AEs (n = 444 solicited local and systemic events across both age cohorts) were rated as mild or moderate, and occurred more frequently in the non-elderly than in the elderly cohort (overall number of events: 247/444 [55.6%] vs. 197/444 [44.4%]), with frequencies increasing in a dose-dependent manner. When fever occurred (n = 18 [18.8%] participants across both age cohorts), it usually resolved within 1 day and was rated predominantly as mild (body temperature 38.0–38.4°C) or moderate (38.5–38.9°C) (**Figure 2**).

**Figure 2.**
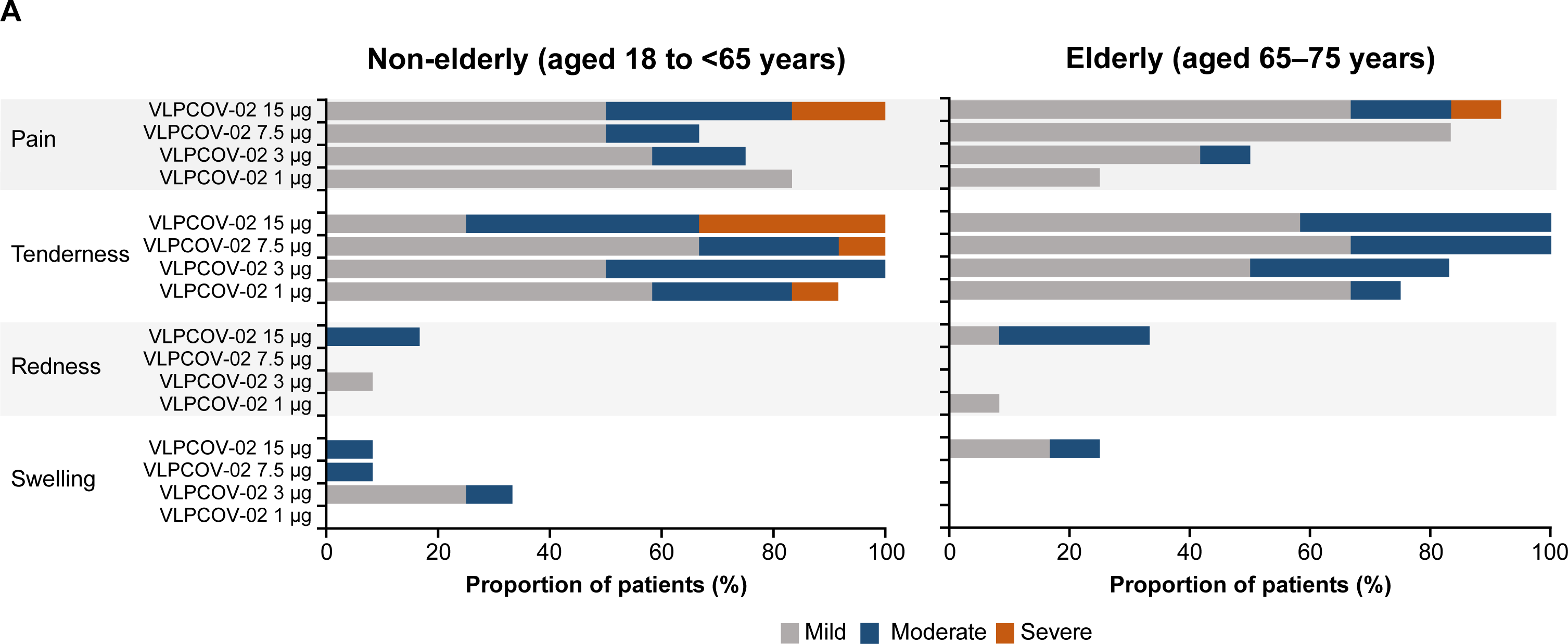
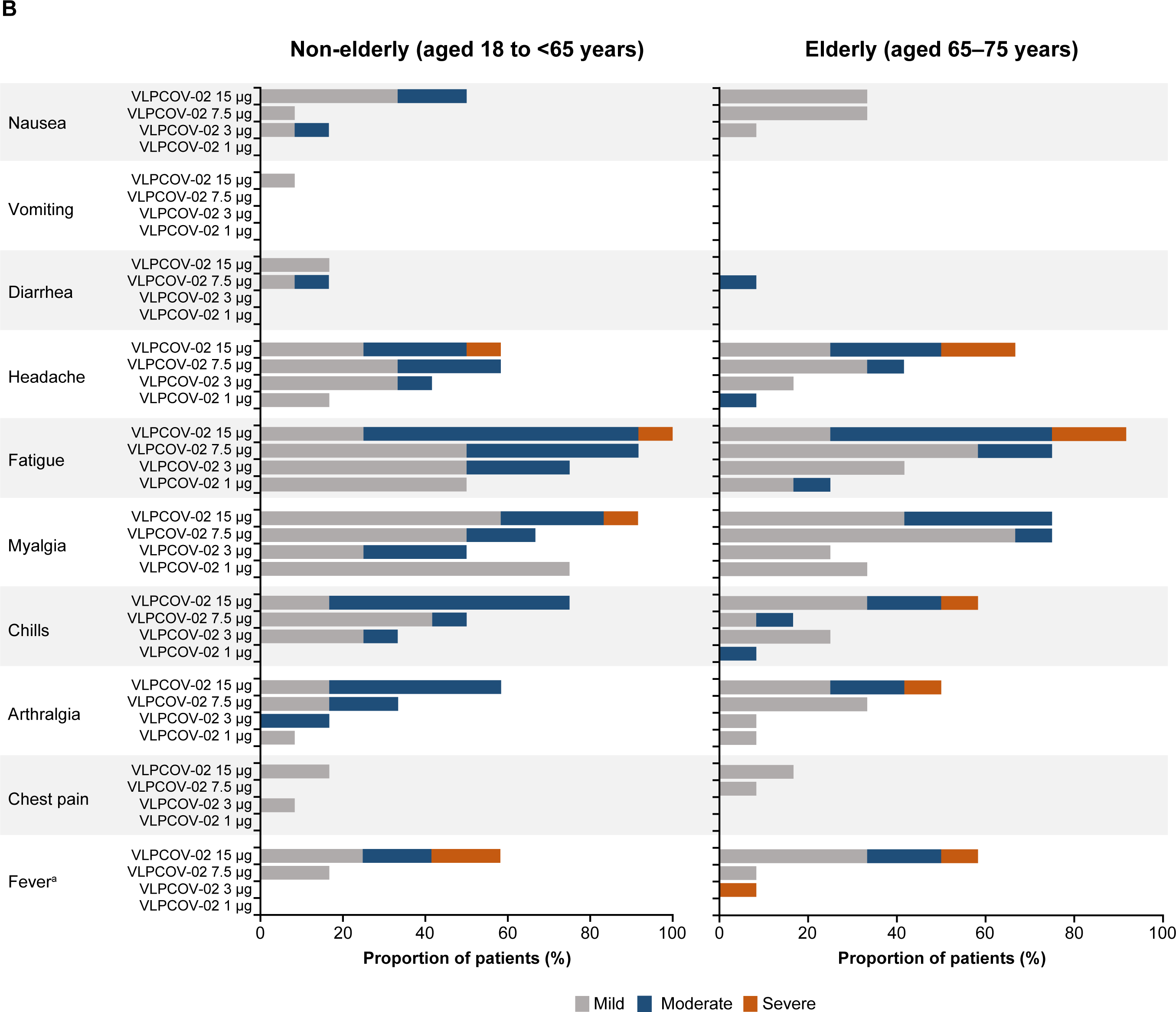
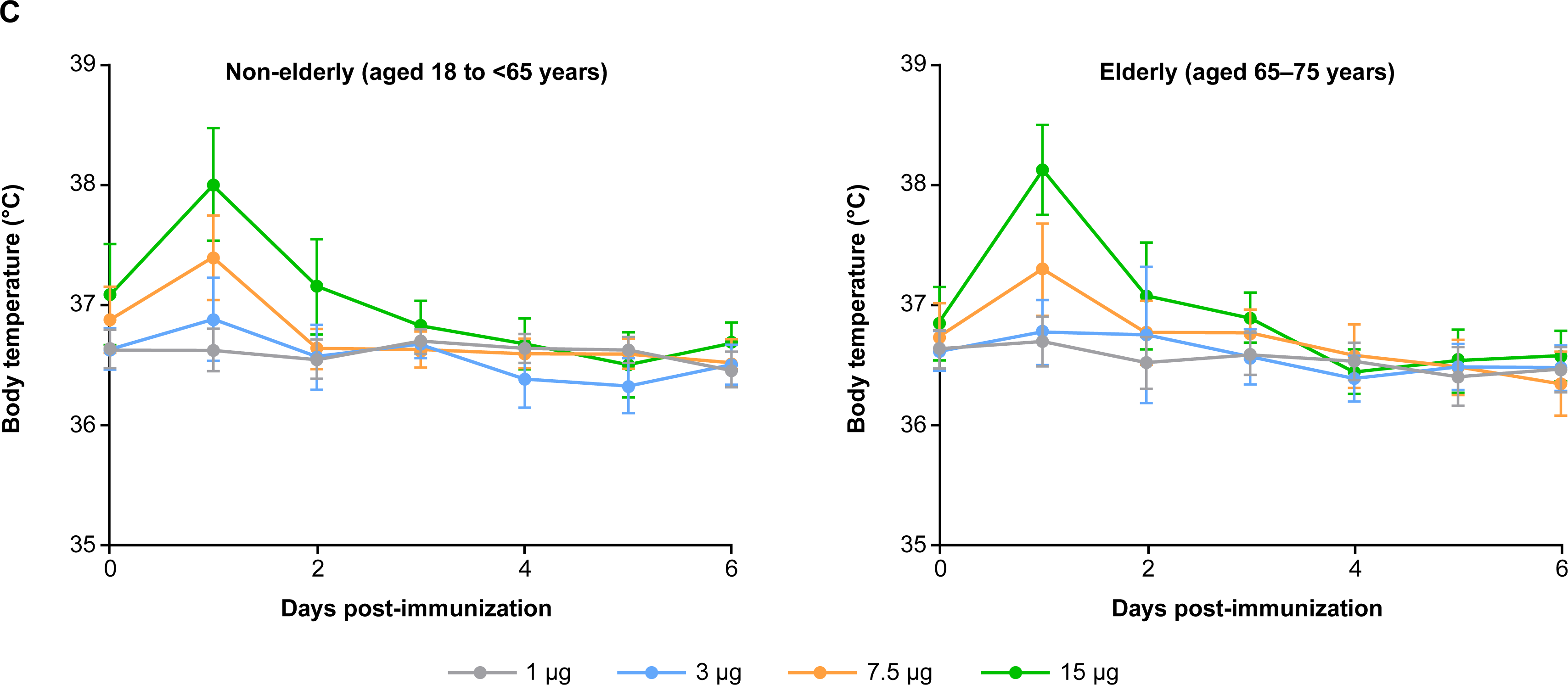
Solicited local and systemic adverse events reported up to 1 week after vaccination with VLPCOV-02. **(A) Solicited local adverse events in the non-elderly and elderly cohorts, by dose level. (B) Solicited systemic adverse events in the non-elderly and elderly cohorts, by dose level. (C) Body temperature (measured orally) in the non-elderly and elderly cohorts, by dose level.** Solicited adverse events were recorded using an E-Diary application. ^a^Fever was graded as follows: mild = 38.0–38.4°C; moderate = 38.5–38.9°C; severe = 39.0–40.0°C.

### Solicited local AEs

The most frequently encountered solicited local AEs across the non-elderly cohort were tenderness (n = 47) and pain (n = 39). Solicited local AEs that were rated as severe were: tenderness, which occurred in one (8.3%) participant who received 1 µg VLPCOV-02, one (8.3%) who received 7.5 µg VLPCOV-02, and four (33.3%) who received 15 µg VLPCOV-02; and pain, which was reported for two (16.7%) participants who received 15 µg VLPCOV-02. Redness occurred in three participants, and was rated as mild for one (8.3%) who received 3 µg VLPCOV-02 and moderate for two (16.7%) who received 15 µg VLPCOV-02. Of note, participants receiving 1 µg VLPCOV-02 experienced no redness or swelling, and any pain (n = 6, 50.0%) was rated as mild (**Figure 2A**).

The most commonly occurring solicited local AEs in the elderly cohort were also tenderness (n = 43) and pain (n = 30). The only solicited local AE rated as severe in this cohort was pain, which was experienced by one (8.3%) participant who received 15 µg VLPCOV-02. The solicited local AE rated as moderate in more than one elderly participant at any dose was tenderness, which occurred in five (41.7%) participants who received 15 µg VLPCOV-02, four (33.3%) who received 7.5 µg VLPCOV-02, and four (33.3%) who received 3 µg VLPCOV-02. None of the participants who received 1 µg VLPCOV-02 experienced swelling, and any pain (n = 3, 25.0%), or redness (n = 1, 8.3%) was rated as mild (**Figure 2A**).

### Solicited systemic AEs

The most frequently experienced solicited systemic AEs in the non-elderly cohort were fatigue (n = 38) and myalgia (n = 34). Solicited systemic AEs that were rated as severe occurred only in participants who received 15 µg VLPCOV-02: headache (n = 1, 8.3%), fatigue (n = 1, 8.3%), and myalgia (n = 1, 8.3%). Participants who received 1 µg VLPCOV-02 reported four of the solicited AEs, all of which were rated as mild: headache (n = 2, 16.7%), fatigue (n = 6, 50.0%), myalgia (n = 9, 75.0%), and arthralgia (n = 1, 8.3%). Mild chest pain occurred in three participants, one (8.3%) who received 3 µg VLPCOV-02 and two who received 15 µg VLPCOV-02. Vomiting was experienced by one (8.3%) non-elderly participant who received 15 µg VLPCOV-02. Fever occurred in nine (18.8%) non-elderly participants, but only at the two highest VLPCOV-02 dose levels: in two participants at 7.5 µg and in seven participants at 15 µg (**Figure 2B**). Two (4.2%) participants, who received 15 µg VLPCOV-02, experienced fever rated as severe (body temperature 39.0–40.0°C); there were no cases of life-threatening fever (i.e., body temperature >40.0°C).

The most commonly encountered solicited systemic AEs reported for the elderly cohort were fatigue (n = 28) and myalgia (n = 25). Solicited systemic AEs rated as severe occurred predominantly in elderly participants who received 15 µg VLPCOV-02: arthralgia (n = 1, 8.3%), chills (n = 1, 8.3%), fever (n = 1, 8.3%), fatigue (n = 2, 16.7%), and headache (n = 2, 16.7%). There were no cases of vomiting, and only one (8.3%) elderly participant reported diarrhea, which was rated as moderate. Mild chest pain occurred in one (8.3%) participant who received 7.5 µg VLPCOV-02 and two (16.7%) who received 15 µg VLPCOV-02. A total of nine (18.8%) fever events were reported for elderly individuals; these events were rated predominantly as mild or moderate and lasted for 1 day (2 days for one participant in the 15 µg group). Severe fever was observed in two participants, one who received 3 µg VLPCOV-02 and another who received 15 µg VLPCOV-02, and was not considered life-threatening (**Figure 2B**).

### Reactogenicity events

The body temperature of all participants who received VLPCOV-02 (N = 96) peaked within 24 h post-immunization, regardless of age cohort or dose of vaccine, and returned to normal within 1 day in all but one participant, in whom it lasted 2 days. The frequency and magnitude of rise in body temperature at 24 hours increased in a dose-dependent manner, and were particularly low in the groups who received lower vaccine doses (1 µg and 3 µg) (**Figure 2C**).

### Immunogenicity

Serum SARS-CoV-2 immunoglobulin (IgG) titers were enhanced during the 4 weeks following VLPCOV-02 booster vaccination across age cohorts (**Figure 3**). Geometric mean titers increased between day 0 and day 28 at each dose level in both the non-elderly and elderly cohorts, by 2.3- to 5.1-fold in the non-elderly cohort and by 2.3- to 2.9-fold in the elderly cohort (**Tables S1–S4**).

**Figure 3.**
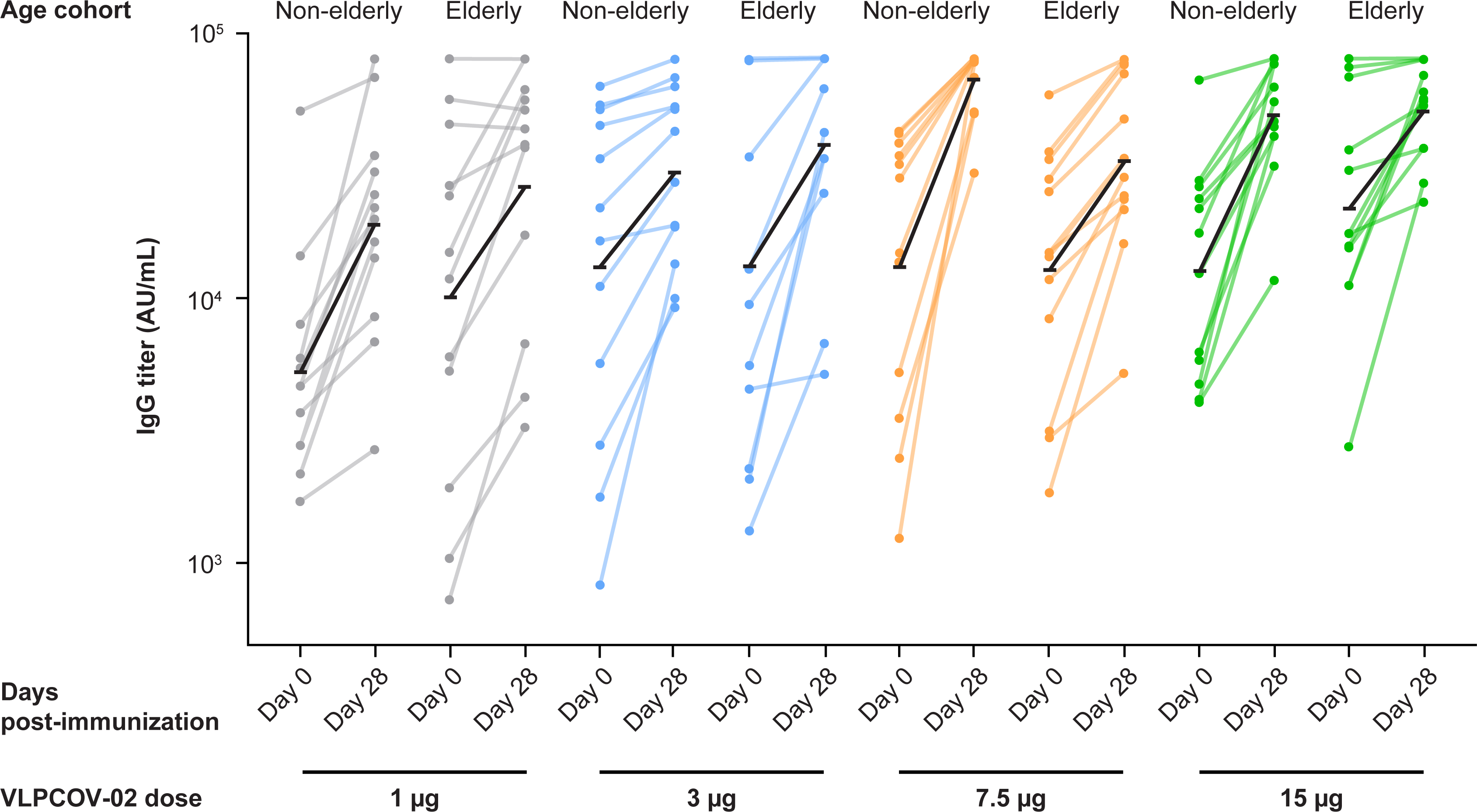
Serum immunoglobulin titers against SARS-CoV-2 RBD protein. Serum SARS-CoV-2 IgG titers against SARS-CoV-2 RBD protein in the non-elderly and elderly cohorts at baseline and at 4 weeks after vaccination, by dose level. The black line indicates the change in geometric mean value of the IgG titers. The maximum titer measurable was 80,000; any values ≥80,000 were treated as 80,000 for the purposes of calculating the geometric mean titers and 95% confidence intervals. IgG, immunoglobulin; RBD, receptor binding domain; SARS-CoV-2, severe acute respiratory syndrome coronavirus 2.

## DISCUSSION

VLPCOV-02 was well tolerated in healthy Japanese adults who had previously received primary vaccination with at least two doses of an approved SARS-CoV-2 vaccine, regardless of their age. VLPCOV-02 was also highly immunogenic, inducing enhancement of serum SARS-CoV-2 IgG titers across all doses tested.

A similar phase 1 study (VLPCOV-01-0102) directly compared booster vaccination with VLPCOV-01 (dosed at 0.3, 1, or 3 µg) with the mRNA BNT162b2 vaccine (30 µg) or placebo (0.9% saline) in 92 healthy Japanese adults (aged 18 to ≥65 years) who had completed two doses of BNT162b2 6–12 months previously.^11^ That study revealed a comparable safety profile between VLPCOV-01 and BNT162b2. No new safety signals were identified with VLPCOV-02 over those reported for VLPCOV-01.^11^ Both vaccines were well tolerated and demonstrated a lower incidence of solicited local and systemic AEs in the elderly compared with the non-elderly cohort, and no unsolicited AEs of concern.^11^ Importantly, the modified vaccine, VLPCOV-02, demonstrated a more favorable safety profile compared with VLPCOV-01, although there has been no direct, head-to-head comparison between the two vaccines. Most solicited AEs for both the VLPCOV-01 and VLPCOV-02 vaccines were rated as mild or moderate. The incidence of severe solicited AEs among VLPCOV-01–vaccinated participants was similar to that for those vaccinated with VLPCOV-02 in the present study (11/324 [3.4%] events vs. 22/480 [4.6%] events). This finding is notable given that half of the population in the present study received a vaccine dose that was double or five times higher than the highest dose of VLPCOV-01 delivered in the phase 1 study (7.5 µg and 15 µg VLPCOV-02 vs. 3 µg VLPCOV-01). Severe solicited systemic AEs were encountered predominantly among participants who received 15 µg VLPCOV-02, regardless of age; only one occurred at the lower doses of VLPCOV-02, 1–3 µg, compared with nine among participants receiving VLPCOV-01 at doses of 0.3–3 µg.^11^

Of particular note, VLPCOV-02 appeared to be less reactogenic than VLPCOV-01, as suggested by comparison of rates of fever in participants receiving the same dose of vaccine. Among the 20 participants who received 3 µg VLPCOV-01, six (30.0%) experienced fever, of whom five were from the non-elderly cohort (n = 10, 50%) and one was from the elderly cohort (n = 10, 10%).^11^ In the present study, among the 24 (non-elderly and elderly) participants who received 3 µg VLPCOV-02, only one (4.2%), an individual in the elderly cohort (n = 12, 8.3%), experienced fever. Notably, the onset of fever in that participant was observed on day 3 after vaccination and lasted for 1 day. None of the 12 participants in the non-elderly cohort experienced fever.

VLPCOV-02 was highly immunogenic, increasing serum SARS-CoV-2 IgG titers during the 4 weeks following vaccination. Baseline IgG titers were higher in the present study compared with the VLPCOV-01 study, a difference that may be attributable to the higher number of vaccinations with publicly available vaccines in the present study population. This observation could also be the result of differences in the eligibility criteria with respect to COVID-19 infection history, and the type and timing of previous vaccines between the VLPCOV-01-0102 and VLPCOV-02-0102 studies. Whilst in VLPCOV-01-0102 individuals were excluded if they had a history of SARS-CoV-2 infection (self-reported and/or confirmatory anti–N-IgG antibody test), in the present VLPCOV-02-0102 study individuals were ineligible if they had previous history of SARS-CoV-2 infection within 6 months of the start of the study, or if they had a positive SARS-CoV-2 antigen test prior to vaccination with VLPCOV-02. In addition, participants could receive vaccination with VLPCOV-01 6–12 months after completion of two doses of the approved novel coronavirus vaccine BNT162b2. For the VLPCOV-02-0102 study, participants could receive VLPCOV-02 vaccination ≥6 months after receiving two doses of the same mRNA vaccine with or without subsequent immunization with any type or number of mRNA vaccines. Thus, in the present study 12 (12.5%) participants were anti–N-IgG positive (i.e., titer ≥1.4), among whom ten (10.4%) had a baseline S protein IgG titer of ≥40,000. The immunogenicity studies with VLPCOV-01 also showed a correlation between anti-RBD IgG titers and pseudovirus-neutralizing responses to VLPCOV-01 booster vaccination. Persistence of IgG titers and associations with neutralizing antibody responses following VLPCOV-02 have yet to be evaluated; these data will be published with analyses from an upcoming study using larger cohort number.

### Study limitations

This study has some limitations, including the small study population, short follow-up duration, open-label design, lack of participants under the age of 35 years, and lack of a comparator arm. Safety reporting for the present study is ongoing, with data planned for collection up to 52 weeks after vaccination.

### Conclusions

The results from this study indicate that the 5-MeC–modified, LNP-encapsulated saRNA VLPCOV-02 vaccine has lower reactogenicity compared with that demonstrated by its predecessor VLPCOV-01, and produces enhanced immune responses. The safety profile also favored VLPCOV-02, with a lower incidence of AEs and fewer occurrences of fever at equivalent doses. VLPCOV-02 may therefore represent an improvement over the non–base-modified VLPCOV-01 as a COVID-19 booster vaccination. Further development of this vaccine is warranted.

## Data Availability

All data produced in the present study are available upon reasonable request to the authors.

## ACKNOWLEDGEMENTS

VLP Therapeutics served as the trial sponsor and was responsible for the design and conduct of the trial, the collection, analysis, and interpretation of the data, and for the writing of the manuscript. This study was supported by AMED under Grant Number JP21nf0101627. Medical writing support was provided by Jacqueline Kolston, PhD (Parexel International) and was funded by VLP Therapeutics, Japan, Inc. All participants provided written informed consent to participate before enrollment.

## Author contributions

M.A., D.K., N.S, S.S., J.F.S, T.S., T.F, Y.I, T.Y, and W.A conceived, designed, and coordinated the study. M.A, D.K, K.K, Y.K, S.S, and N.S prepared and executed the clinical study. K.M, D.K, K.K, Y.K, M.K, A.L.M, M.A., and W.A. contributed to the data validation, analysis, writing, and revision of the manuscript. All authors read and approved the manuscript, had full access to all the data in the study, and had final responsibility for the decision to submit for publication.

## Experimental model and subject details

### Method details

#### Study design and population

VLPCOV-02-0102 is an ongoing, two-part phase 1/2 study. Part 1 is a phase 1 dose-escalation, open-label study to assess the safety and tolerability of VLPCOV-02 as a single booster dose in healthy individuals who have received primary vaccination series with approved COVID-19 vaccines. Part 2 is a phase 2 multicenter, randomized, active comparator-controlled, observer-blinded study to determine the recommended booster dose. The findings of Part 1 up to 4 weeks after vaccination are presented here.

Eligible participants for Part 1 were healthy Japanese adults (age ≥18–75 years at study entry) at the Medical Corporation OPHAC Hospital in Japan who had received the two-dose primary vaccination series with the same COVID-19 uridine-modified RNA vaccine (hereafter “mRNA vaccine”) or a booster vaccination with an mRNA vaccine (any kind, any number, and including bivalent mRNA vaccines) ≥6 months previously. Key exclusion criteria included a history of COVID-19 ≤6 months prior to study day 1, persistent symptoms of any kind following either COVID-19 or previous vaccination against COVID-19; history or presence of a serious cardiovascular, hematologic, respiratory, hepatic, renal, gastrointestinal, and/or neuropsychiatric disease; and presence or known history of a disease, or previous/planned receipt of any agent or therapy that could affect immunogenicity assessments. Pregnant and lactating females were also excluded from study entry.

#### Trial procedures

The participants were grouped according to age: non-elderly (age 18 to <65 years) and elderly (age 65–75 years). Four dose levels of VLPCOV-02 were planned for sequential testing in both age cohorts: 1, 3, 7.5, and 15 μg. All vaccinations were delivered as single, intramuscular injections to the upper-arm deltoid muscle of each participant, who were then carefully monitored for ≥30 min after administration for assessment of reactogenicity. Medical interview, phonacoscopy, blood pressure, pulse rate, body temperature (measured orally), and 12-lead ECG were performed before and 2 h after administration. Follow-up visits were scheduled from day 4 up to week 52 after vaccination to identify any changes in concomitant medications, collect vital signs, review AEs, and obtain blood samples for immunogenicity analyses.

Transition from (1) the sentinel participant at each dose level to subsequent participants at the same dose level and (2) the sentinel participant in the non-elderly cohort to their counterpart in the elderly cohort at the same dose level was confirmed after the principal investigator determined there were no safety concerns (**Figure 1**). Transition from one VLPCOV-02 dose level to the next higher dose level within the same age cohort proceeded after the principal investigator (upon discussion of prespecified criteria with the sponsor and medical expert) determined that there were no medical concerns up to day 4 after vaccination. If progression to the higher dose was not allowed, the dose was reduced (**Figure 1**).

#### Safety assessments

The primary safety endpoints were incidence and severity (rated as mild, moderate, or severe) of solicited local (pain, tenderness, redness, induration, and swelling) and systemic (nausea, vomiting, diarrhea, headache, fatigue, myalgia, chills, chest pain, and fever) AEs occurring up to 1 week after vaccination, and any AEs occurring up to 4 weeks after vaccination. Incidence of serious AEs, COVID-19, and AEs leading to study discontinuation would be captured for up to 52 weeks after vaccination or until study discontinuation. All other AEs were assessed until 4 weeks after vaccination. If symptoms of suspected myocarditis/pericarditis were recorded, 12-lead ECG and blood biochemistry (creatine kinase, C-reactive protein, cardiac troponin I) were conducted, with diagnostic imaging as needed. The severity of most AEs was assessed using Common Terminology Criteria in Solid Tumors version 5.0 as well as per United States Food and Drug Administration guidance (Toxicity Grading Scale for Healthy Adult and Adolescent Volunteers Enrolled in Preventive Clinical Trials).^19^ The severity of fever was rated as mild, moderate, or severe at body temperatures (measured orally) of 38.0–38.4°C, 38.5–38.9°C and 39.0–40.0°C, respectively, and was considered life-threatening at body temperatures >40.0°C.

#### Immunogenicity assessments

The primary immunogenicity endpoints were geometric mean titer (GMT) and seroresponse rate of serum IgG titers against SARS-CoV-2 RBD protein at 4 weeks after vaccination. Only GMT was calculated for this interim analysis. Serum IgG titers were quantified in samples analyzed using the SARS-CoV-2 IgG II Quant assay, which detects IgG antibodies to the RBD of the SARS-CoV-2 spike protein, according to the manufacturer’s instructions (maximum titer measurable, 80,000; Abbott Laboratories).

#### Statistical analysis

The data cutoff date for this interim analysis, which was not prespecified, was June 18, 2023. The planned study enrollment was 96 participants (48 non-elderly and 48 elderly), with each dose level including 12 participants (one sentinel and 11 subsequent participants; no sentinel participant was required in the case where the dose level was reduced). The safety analysis population included all participants who received the booster vaccination with VLPCOV-02 (at any dose). Safety analyses included in this report were analyzed descriptively and are presented as numbers and percentages of participants who experienced solicited AEs (local and systemic) up to 1 week after vaccination, AEs up to 4 weeks after vaccination, and serious AEs, COVID-19, and AEs leading to study discontinuation up to 4 weeks after vaccination. GMTs of serum IgG titers against SARS-CoV-2 RBD were calculated by group at each measurement time point. In the case where participants experienced COVID-19 infection during the study, data points that were impacted (i.e., recorded during COVID-19 infection) were excluded from the analyses.

### Additional resources

The study protocol and any amendments were approved by Nobuaki Sato (VLP Therapeutics Japan, Inc.) and complied with relevant regulatory requirements. This trial is registered with the Japan Registry of Clinical Trials (jRCT2051230005) and the Provincial Disaster Management Authority (2022-8076).

## SUPPLEMENT

### SUPPLEMENTARY RESULTS

**Figure S1.**
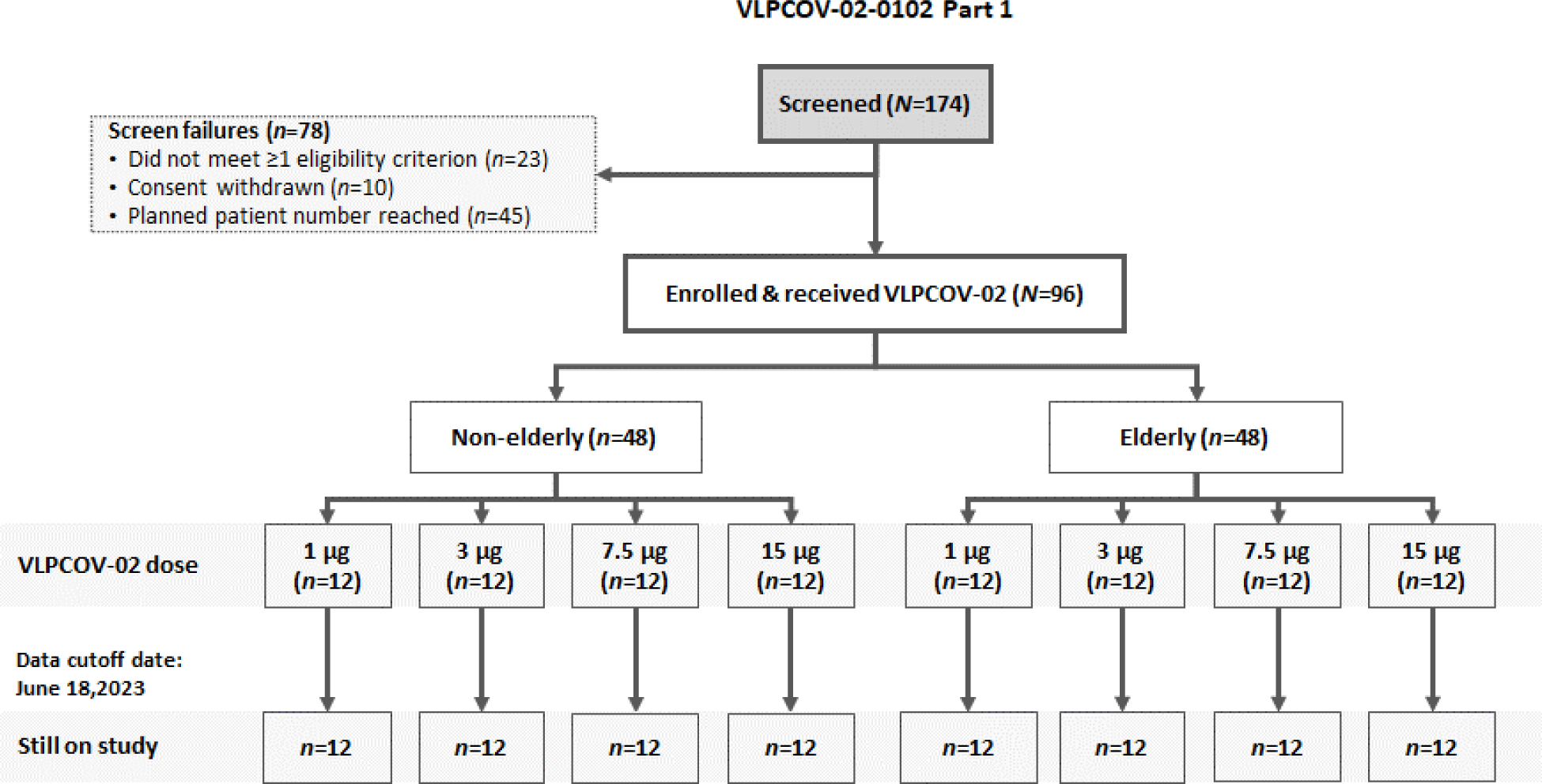
Study disposition flow diagram.

**Table S1.**
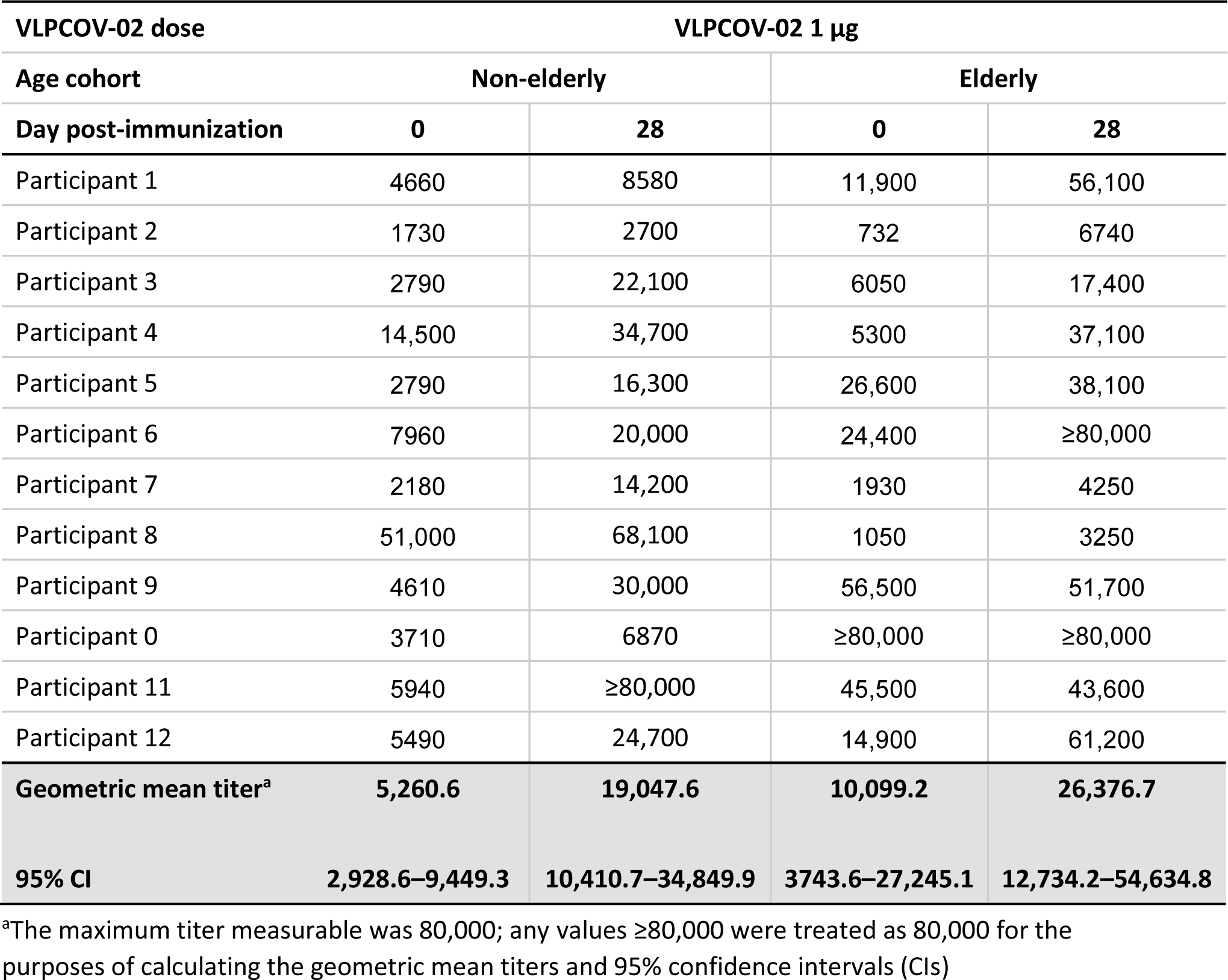
Serum SARS-CoV-2 IgG titers^a^ at baseline (day 0) and 4 weeks after vaccination (day 28) by participant, and geometric mean titer for participants in the non-elderly and elderly cohorts who received VLPCOV-02 1 µg.

**Table S2.** Serum SARS-CoV-2 IgG titers^a^ at baseline (day 0) and 4 weeks after vaccination (day 28) by participant, and geometric mean titer for participants in the non-elderly and elderly cohorts who received VLPCOV-02 3 µg.

| VLPCOV-02 dose |  | VLPCOV-02 3 µg |  |  |
| --- | --- | --- | --- | --- |
| Age cohort | Non-elderly |  | Elderly |  |
| Day post-immunization | 0 | 28 | 0 | 28 |
| Participant 1 | 51,600 | 68,000 | ≥80,000 | ≥80,000 |
| Participant 2 | 835 | 13,600 | 2290 | 33,700 |
| Participant 3 | 16,500 | 18,900 | 13,000 | 61,800 |
| Participant 4 | 21,900 | 42,800 | 5620 | 33,300 |
| Participant 5 | 33,600 | 52,200 | 9530 | 25,000 |
| Participant 6 | 11,100 | 27,600 | ≥80,000 | ≥80,000 |
| Participant 7 | 62,900 | ≥80,000 | 78,500 | ≥80,000 |
| Participant 8 | 2800 | 9270 | 2090 | 42,400 |
| Participant 9 | 44,800 | 53,200 | 1330 | 6770 |
| Participant 0 | 1780 | 10,000 | ≥80,000 | ≥80,000 |
| Participant 11 | 5690 | 18,700 | 4560 | 5200 |
| Participant 12 | 53,400 | 63,200 | 34,100 | ≥80,000 |
| **Geometric mean titer<sup>a</sup>** | **13,066.9** | **29,860.6** | **13,188.8** | **37,789.2** |
| **95% CI** | **5142.3–33,204.0** | **18,251.4–48,853.8** | **4828.1–36,027.4** | **20,521.1–69,588.0** |

<sup>a</sup>The maximum titer measurable was 80,000; any values ≥80,000 were treated as 80,000 for the purposes of calculating the geometric mean titers and 95% confidence intervals (CIs)

**Table S3.** Serum SARS-CoV-2 IgG titers^a^ at baseline (day 0) and 4 weeks after vaccination (day 28) by participant, and geometric mean titer for participants in the non-elderly and elderly cohorts who received VLPCOV-02 7.5 µg.

| VLPCOV-02 dose |  | VLPCOV-02 7.5 µg |  |  |
| --- | --- | --- | --- | --- |
| Age cohort | Non-elderly |  | Elderly |  |
| Day post-immunization | 0 | 28 | 0 | 28 |
| Participant 1 | 38,600 | 77,300 | 14,500 | 33,900 |
| Participant 2 | 5260 | 49,300 | 58,800 | ≥80,000 |
| Participant 3 | 41,700 | ≥80,000 | 3140 | 23,700 |
| Participant 4 | 13,700 | 67,800 | 1860 | 16,100 |
| Participant 5 | 14,900 | ≥80,000 | 25,300 | 47,400 |
| Participant 6 | 42,800 | ≥80,000 | 8400 | 28,800 |
| Participant 7 | 32,200 | ≥80,000 | 33,500 | 76,600 |
| Participant 8 | 28,500 | ≥80,000 | 28,300 | 70,500 |
| Participant 9 | 2500 | 50,700 | 11,800 | 21,800 |
| Participant 0 | 34,700 | ≥80,000 | 2990 | 5230 |
| Participant 11 | 3540 | 29,600 | 35,700 | ≥80,000 |
| Participant 12 | 1260 | ≥80,000 | 14,900 | 24,500 |
| **Geometric mean titer<sup>a</sup>** | **13,130.6** | **66,965.1** | **12,771.3** | **33,060.2** |
| **95% CI** | **5951.5–28,969.3** | **54,877.1–81,715.9** | **6329.3–25,770.1** | **19,714.5–55,440.4** |

<sup>a</sup>The maximum titer measurable was 80,000; any values ≥80,000 were treated as 80,000 for the purposes of calculating the geometric mean titers and 95% confidence intervals (CIs)

**Table S4.**
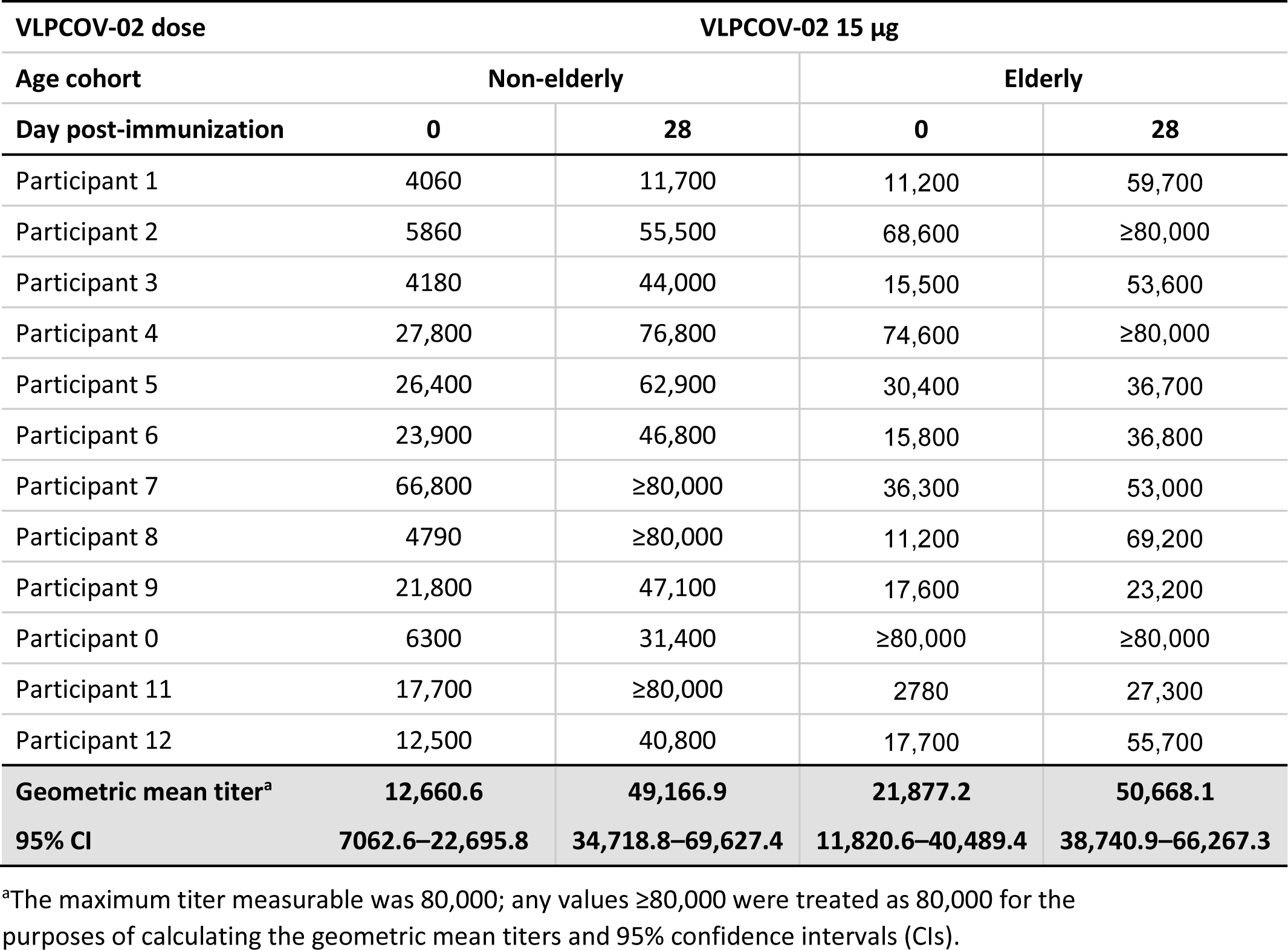
Serum SARS-CoV-2 IgG titers^a^ at baseline (day 0) and 4 weeks after vaccination (day 28) by participant, and geometric mean titer for participants in the non-elderly and elderly cohorts who received VLPCOV-02 15 µg.

| VLPCOV-02 dose |  | VLPCOV-02 15 µg |  |  |
| --- | --- | --- | --- | --- |
| Age cohort | Non-elderly |  | Elderly |  |
| Day post-immunization | 0 | 28 | 0 | 28 |
| Participant 1 | 4060 | 11,700 | 11,200 | 59,700 |
| Participant 2 | 5860 | 55,500 | 68,600 | ≥80,000 |
| Participant 3 | 4180 | 44,000 | 15,500 | 53,600 |
| Participant 4 | 27,800 | 76,800 | 74,600 | ≥80,000 |
| Participant 5 | 26,400 | 62,900 | 30,400 | 36,700 |
| Participant 6 | 23,900 | 46,800 | 15,800 | 36,800 |
| Participant 7 | 66,800 | ≥80,000 | 36,300 | 53,000 |
| Participant 8 | 4790 | ≥80,000 | 11,200 | 69,200 |
| Participant 9 | 21,800 | 47,100 | 17,600 | 23,200 |
| Participant 0 | 6300 | 31,400 | ≥80,000 | ≥80,000 |
| Participant 11 | 17,700 | ≥80,000 | 2780 | 27,300 |
| Participant 12 | 12,500 | 40,800 | 17,700 | 55,700 |
| **Geometric mean titer<sup>a</sup>** | **12,660.6** | **49,166.9** | **21,877.2** | **50,668.1** |
| **95% CI** | **7062.6–22,695.8** | **34,718.8–69,627.4** | **11,820.6–40,489.4** | **38,740.9–66,267.3** |

<sup>a</sup>The maximum titer measurable was 80,000; any values ≥80,000 were treated as 80,000 for the purposes of calculating the geometric mean titers and 95% confidence intervals (CIs).

